# AI-based identification of patients who benefit from revascularization: a multicenter study

**DOI:** 10.1101/2025.06.11.25329295

**Authors:** Wenhao Zhang, Robert JH Miller, Krishna Patel, Aakash D. Shanbhag, Joanna X Liang, Mark Lemley, Giselle Ramirez, Valerie Builoff, Jirong Yi, Jianhang Zhou, Paul Kavanagh, Wanda Acampa, Timothy M Bateman, Marcelo Di Carli, Sharmila Dorbala, Andrew J Einstein, Mathews B Fish, M. Timothy Hauser, Terrence D. Ruddy, Philipp A Kaufmann, Edward J Miller, Tali Sharir, Monica Martins, Julian Halcox, Panithaya Chareonthaitawee, Damini Dey, Daniel S Berman, Piotr J Slomka

## Abstract

**Background and Aims:** Revascularization in stable coronary artery disease often relies on ischemia severity, but we introduce an AI-driven approach that uses clinical and imaging data to estimate individualized treatment effects and guide personalized decisions.

**Methods:** Using a large, international registry from 13 centers, we developed an AI model to estimate individual treatment effects by simulating outcomes under alternative therapeutic strategies. The model was trained on an internal cohort constructed using 1:1 propensity score matching to emulate randomized controlled trials (RCTs), creating balanced patient pairs in which only the treatment strategy—early revascularization (defined as any procedure within 90 days of MPI) versus medical therapy—differed. This design allowed the model to estimate individualized treatment effects, forming the basis for counterfactual reasoning at the patient level. We then derived the AI-REVASC score, which quantifies the potential benefit, for each patient, of early revascularization. The score was validated in the held-out testing cohort using Cox regression.

**Results:** Of 45,252 patients, 19,935 (44.1%) were female, median age 65 (IQR: 57-73). During a median follow-up of 3.6 years (IQR: 2.7-4.9), 4,323 (9.6%) experienced MI or death. The AI model identified a group (n=1,335, 5.9%) that benefits from early revascularization with a propensity-adjusted hazard ratio of 0.50 (95% CI: 0.25-1.00). Patients identified for early revascularization had higher prevalence of hypertension, diabetes, dyslipidemia, and lower LVEF.

**Conclusions:** This study pioneers a scalable, data-driven approach that emulates randomized trials using retrospective data. The AI-REVASC score enables precision revascularization decisions where guidelines and RCTs fall short.

**Graphical Abstract:** 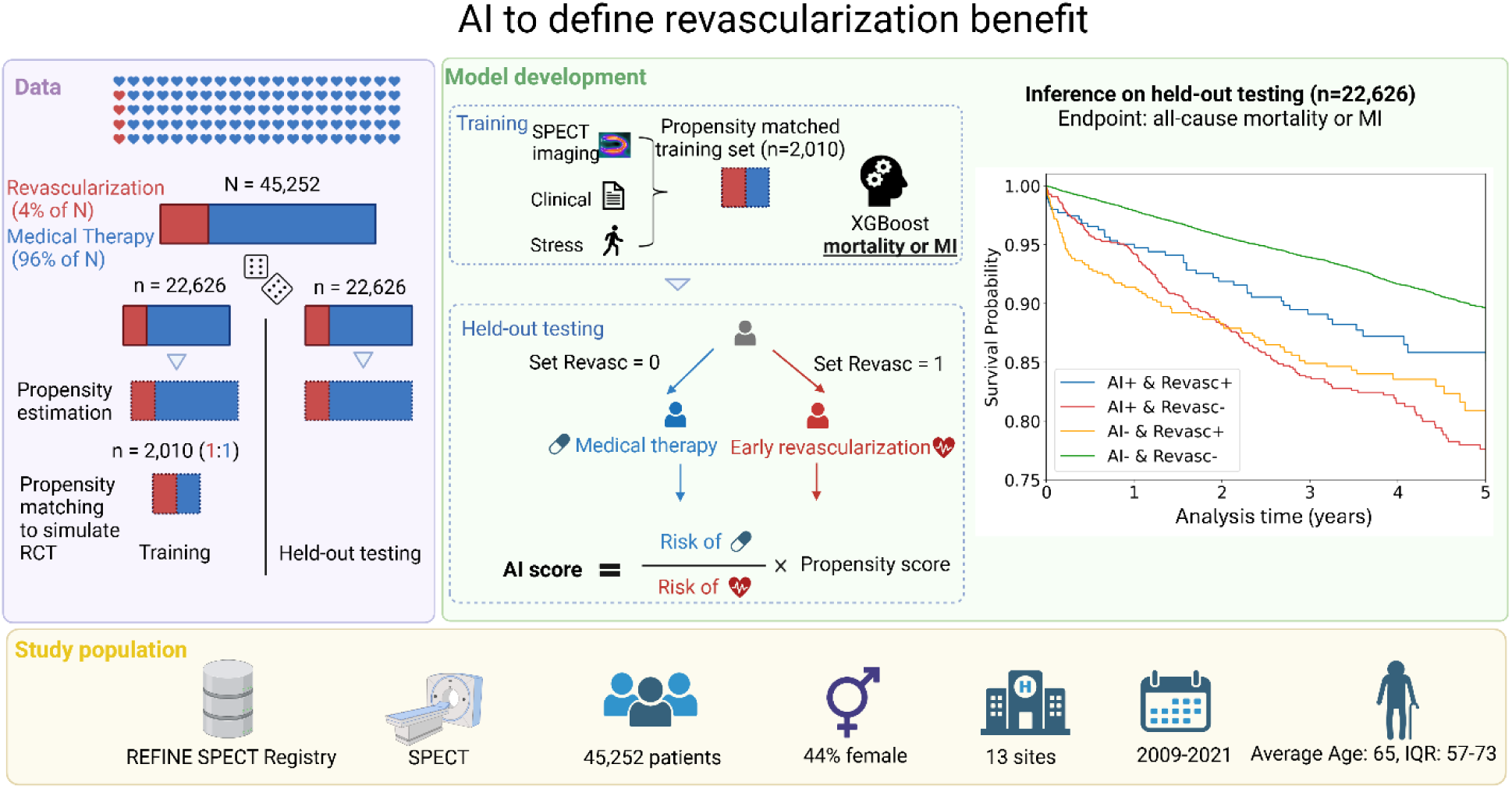

## 1. INTRODUCTION

Myocardial perfusion imaging (MPI) is frequently performed to guide therapy in patients with known or suspected coronary artery disease (CAD)^1^. While most patients can be managed medically^2^, patients with symptoms due to ischemia may derive benefit from revascularization^3^. It has not yet been demonstrated that patients with chronic coronary syndromes who derive a survival benefit from revascularization can be reliably identified. Prior observational studies have suggested that myocardial ischemia could identify patients who may benefit from revascularization ^4–9^. However, the International Study of Comparative Health Effectiveness with Medical and Invasive Approaches (ISCHEMIA) trial did not confirm this hypothesis^10, 11^, suggesting that methods incorporating additional factors are needed to provide optimal, patient-specific selection of revascularization.

Recent observational studies have demonstrated potential variability across subgroups which could help improve selection of patients for revascularization. For example, survival benefit from early revascularization may be impacted by the presence of left ventricular dysfunction^7^, which has also been suggested by sub-group analysis of the ISCHEMIA trial^2^. Similarly, the presence of diabetes may influence the ischemia threshold at which patients benefit from revascularization^12^. These nuanced interactions among clinical and imaging variables are difficult to apply consistently in clinical practice. While artificial intelligence (AI) methods integrating clinical and imaging variables have been applied to risk stratification in prior studies^13^, the key clinical challenge remains: identifying which patients experience the greatest benefit from early revascularization rather than simply identifying those at the highest risk.

Here, we propose and validate a novel AI-based approach (AI-REVASC score) that integrates clinical and imaging features to guide individualized revascularization decisions following myocardial perfusion imaging (MPI). By combining counterfactual reasoning and a digital twin approach—simulating how a specific patient might have responded to an alternative treatment path—we can estimate individual treatment effects while addressing selection bias in retrospective data. This approach leverages a large, international, multicenter registry—data that would otherwise be rarely available from randomized controlled trials (RCTs), which are often constrained by logistical, and economic considerations that preclude the exhaustive testing of all potential treatment strategies in all patient types. This work pioneers a pathway for precision medicine in cardiology when RCTs are not feasible, offering a scalable, data-driven AI framework for deriving high-quality evidence.

## 2. METHODS

### 2.1 Study Design and Patient Population

We conducted an observational study in patients with known or suspected CAD to assess the potential benefit of early revascularization in relation to burden of inducible ischemia. A total of 45,252 consecutive patients from 13 centers from the United States, Canada, Switzerland, Israel, and Italy who underwent MPI were included in the study^14, 15^. The cohort included consecutive patients at each center referred for single photon emission computed tomography (SPECT) MPI between 2009 and 2021. Patients were imaged with both solid-state and conventional camera systems^15^. The study protocol complied with the Declaration of Helsinki and was approved by the institutional review boards at each participating institution. The investigators ensured that the institutional ethics committee at each center evaluated and approved the study protocol before data collection and transfer. The overall study was approved by the institutional review board at Cedars-Sinai Medical Center (Office of Research Compliance and Quality Improvement). Sites either obtained written informed consent or waiver of consent for the use of the de-identified data.

The population was divided into 2 groups: patients who underwent early revascularization, defined as percutaneous coronary intervention (PCI) or coronary artery bypass graft surgery (CABG) within 90 days after MPI^4–7^, and those who were initially treated medically.

### 2.2 Clinical Data

Baseline demographic information included age, sex, family history of premature CAD, smoking status, history of hypertension, hyperlipidemia, diabetes, or peripheral vascular disease and previous myocardial infarction (MI), PCI or CABG and chest pain prior to testing. Stress types were classified as exercise or pharmacologic and stress electrocardiogram (ECG) response was interpreted by experienced cardiologists at the time of MPI.

### 2.3 Image Quantification

MPI studies were conducted with multiple camera systems. All imaging data was checked by experienced technologists at the core laboratory (Cedars-Sinai Medical Center, Los Angeles). After quality control, images were quantified with Quantitative Perfusion SPECT (QPS) software for all patients sequentially in an automated batch mode, which optimizes the computational resources required to process the image registry and records all quantitative data automatically for further analysis^16^. All quantitative imaging data, including total perfusion deficit (TPD), stress and rest left ventricle ejection fraction (LVEF), and morphologic measurements were available for the propensity score and AI models.

### 2.4 Visual perfusion assessment

SPECT MPI studies were evaluated visually by experienced physicians at each site during clinical reporting, with access to all relevant data, including stress and rest perfusion imaging, gated functional data, quantitative measurements, and clinical information. The final visual assessment was based on ≥10% myocardial ischemia following the 17-segment model established by the American Heart Association. If clinical scores were not available from the site, quantitative segmental scores were used. ≥10% myocardial ischemia of the myocardium was considered the current clinical decision criterion for early revascularization selection based on ischemia alone. Ischemia-based selection was used as the comparison to AI-based selection as it was utilized for patient selection in the ISCHEMIA trial^10, 11^.

### 2.5 Outcomes

The primary outcome was death or MI over a 5-year period. All-cause mortality was determined from administrative databases which varied by site. MI were adjudicated by experienced cardiologists at each site after considering all available clinical information including symptoms, ECG changes, cardiac biomarkers, non-invasive testing results and invasive angiography.

### 2.6 AI Revascularization Benefit Score

Data was randomly and evenly split into the internal training or held-out testing groups, stratified by all-cause mortality or MI. The internal training set was then further split into a matched cohort using 1:1 nearest neighbor propensity score matching on early revascularization, to emulate the balance of baseline characteristics found in randomized controlled trials.

Individuals not included in the matched cohort were excluded from model development to preserve internal validity and reduce bias due to confounding. The internal dataset included n=2,010 patients. The held-out dataset included n=22,626 patients.

Extreme Gradient Boosting (XGBoost) model was trained on the propensity-matched dataset to predict all-cause mortality or MI over five years, mimicking a randomized controlled trial to reduce confounding and estimate heterogeneous treatment effects. We utilized XGBoost’s default method for missing value imputation since it provides similar prediction performance to more complicated methods for value imputation^17^. Supplementary Figure 1 summarizes the proportion of missing values in the internal training. During model training, 10-fold cross-validation was applied to the internal dataset for model hyper-parameter tuning, using a grid search method aimed to optimize validation loss. The model with the lowest validation loss was applied to the held-out testing population.

During the model inferencing phase, to estimate individualized treatment effects, a digital twin approach was used to simulate each patient’s counterfactual outcome—reversing only the treatment while holding all other characteristics constant. This emulation of an identical patient receiving the alternative therapy enabled comparisons that would be impossible in real-world settings, where only one treatment pathway is observed. The digital twin leveraged the XGBoost model’s RCT-like estimates of treatment effects to generate counterfactual predictions for each patient’s alternative treatment arm. An AI score was calculated for each patient based on the risk reduction when switching from medical therapy to early revascularization. Patients were then ranked by the AI-generated score, with higher scores indicating greater expected benefit from early revascularization.

To define a subgroup predicted to benefit from revascularization, we selected for the AI score, the same percentage of top-scoring patients based on the AI score as the proportion of patients with ischemia ≥10%. Thus, the AI-selected revascularization group matched the size of the ischemia-based selection group.

The model incorporated various characteristics, including clinical and imaging factors. Clinical characteristics encompassed age, gender, and past medical history, smoking, peripheral vascular disease, family history of CAD, past MI, early revascularization, and chest pain symptoms. MPI variables included the type of stress test performed, as well as blood pressure measurements taken during both stress and rest conditions. MPI characteristics consisted of ischemic TPD, rest TPD, ejection fraction, shape index, volume measurements of the left ventricle, and transient ischemic dilation ratio. Note that the model excluded the follow-up duration for mortality, as it is directly tied to the outcome being predicted.

### 2.7 Model Explainability

SHapley Additive exPlanations (SHAP), a game-theoretic feature importance method, was used to explain how features contributed to the overall prediction for individual patients^5^. In the SHAP plot, each dot represents a single patient’s SHAP value for a feature, showing the extent to which each feature drives the prediction for that instance: positive SHAP values indicate an increase in the prediction, while negative values indicate a decrease. The distance from zero on the x-axis reflects the feature’s influence magnitude.

### 2.8 Propensity Score

We utilized propensity score matching to account for non-randomization of revascularization^18^. A logistic regression model was trained only on the training set, and generated propensity scores for internal training set. The propensity scores included clinical characteristics (age, sex, and past medical history), stress test characteristics (mode of stress, stress and rest heart rate and blood pressures), and imaging characteristics (ischemic TPD, rest TPD, stress ejection fraction, shape index, left ventricle volume, and transient ischemic dilation). In the training population, we utilized 1:1 nearest neighbor propensity score matching to ensure equal representation of the early revascularization and medical therapy groups^19, 20^. The covariate balancing of training set by propensity scores were shown in Supplementary Figure 221.

### 2.9 Statistical Analysis

For continuous variables following a normal distribution, data are reported as mean ± standard deviation (SD), while those not normally distributed are presented as medians with interquartile range (IQR) [IQ1-IQ3]. Categorical variables are summarized using counts and relative frequencies (%). Pearson’s χ2 test was used to compare categorical variables, whereas the Wilcoxon Mann-Whitney test was applied for continuous variables when appropriate. Model performance was assessed through receiver-operating characteristic (ROC) analysis, with area under the curve (AUC) values and their 95% confidence intervals (CI) compared using the paired DeLong test^22^. To evaluate associations with the primary outcome, Kaplan-Meier survival curves and univariate Cox proportional hazard models were utilized. A multivariable Cox proportional hazards model was used to evaluate the association of early revascularization with all-cause mortality or MI. Hazard ratios were adjusted by adjusted by clinical characteristics (age, sex, and past medical history), stress test characteristics (mode of stress, stress and rest heart rate and blood pressures), and imaging characteristics (ischemic TPD, rest TPD, stress ejection fraction, shape index, left ventricle volume, and transient ischemic dilation). Statistical significance was determined via the log-rank test, and confidence intervals were estimated using the percentile bootstrap method. A two-tailed p-value of <0.05 was considered statistically significant.

Statistical analyses were performed with Pandas (version 2.1.1) and Numpy (version 1.24.3), Scipy (version 1.11.4), Lifelines (version 0.28.0) and Scikit-learn (version 1.3.0) in Python version 3.11.5 (Python Software Foundation, Wilmington, DE, USA), as well as “nricens” package (version 1.6) in R version 4.3.2 (R Foundation for Statistical Computing, Vienna, Austria).

## 3. RESULTS

### 3.1 Patient Characteristics

Patient characteristics stratified by initial treatment strategy are summarized in Table 1. Among the 45,252 participants, 19,935 (44.1%) were female, with a median age of 65 years (IQR 57–73). Of these, 1,963 (4.3%) underwent early revascularization, while the remaining patients received medical therapy. Those who underwent an invasive strategy were significantly older (66.5 vs. 65.0 years, p<0.001) and had a higher proportion of male patients (76.7% vs 55.0%, p<0.001), patients with diabetes mellitus (37.5% vs. 26.7%, p<0.001), and patients with hypertension (72.9% vs. 63.5%, p<0.001). Additionally, they had lower stress LVEF (56.9 vs. 64.2, p<0.001) and greater ischemic TPD (7.2 vs. 2.0, p<0.001) compared to those who received medical therapy.

**Table 1.** Baseline patient characteristics by treatment.

|  | **Overall**<br>**N = 45,252** | **Early**<br>**Revascularization**<br>**n = 1,963 (4.3%)** | **Medical Therapy**<br>**n=43,289 (95.7%)** | **p-value** |
| --- | --- | --- | --- | --- |
| Age, median (IQR) | 65.0<br>(57.0, 73.0) | 66.5<br>(59.0, 74.0) | 65.0<br>(57.0, 73.0) | <0.001 |
| Male, n (%) | 25,317 (55.9%) | 1,505 (76.7%) | 23,812 (55.0%) | <0.001 |
| BMI, median (IQR) | 28.4<br>(25.1, 32.6) | 28.4<br>(25.5, 32.3) | 28.3<br>(25.1, 32.6) | 0.5 |
| Race, n (%) |  |  |  | <0.001 |
| American Indian or Alaska Native | 143 (0.3%) | 8 (0.4%) | 135 (0.7%) |  |
| Asian | 516 (1.2%) | 25 (1.3%) | 491 (2.7%) |  |
| Black or African American | 2,509 (5.5%) | 69 (3.5%) | 2,440 (13.5%) |  |
| Native Hawaiian or Other Pacific Islander | 41 (0.1%) | 1 (0.0%) | 40 (0.2%) |  |
| White | 13,894 (30.7%) | 655 (33.4%) | 13,239 (73.5%) |  |
| Not available | 28,149 (62.2%) | 1,205 (61.4%) | 26,944 (62.2%) |  |
| **Past Medical History** |  |  |  |  |
| Past CAD, n (%) | 5,272 (11.7%) | 393 (20.0%) | 4,879 (11.3%) | <0.001 |
| Hypertension, n (%) | 28,934 (63.9%) | 1,431 (72.9%) | 27,503 (63.5%) | <0.001 |
| Diabetes Mellitus, n (%) | 12,277 (27.1%) | 736 (37.5%) | 11,541(26.7%) | < 0.001 |
| Hyperlipidemia, n (%) | 23,053 (50.9%) | 1,146 (58.4%) | 27,503 (63.5%) | <0.001 |
| Family History of CAD, n (%) | 14,229 (31.4%) | 677 (34.5%) | 13,552 (31.3%) | 0.004 |
| Peripheral Vascular Disease, n (%) | 8,651 (19.1%) | 654 (33.3%) | 7,997 (18.5%) | <0.001 |
| Smoking, n (%) | 10,841 (24.0%) | 543 (27.7%) | 10,298 (23.8%) | <0.001 |
| Typical chest pain, n (%) | 2,880 (6.4%) | 309 (15.7%) | 2,571 (5.9%) | <0.001 |
| Atypical chest pain, n (%) | 12,331 (27.3%) | 584 (29.8%) | 11,747 (27.1%) | 0.012 |
| MPI Acquisition Parameters |  |  |  |  |
| Exercise Stress, n (%) | 21,016 (46.4%) | 825 (42.0%) | 20,191 (46.6%) | <0.001 |
| Resting Heart Rate, median (IQR) | 71 (62, 81) | 71 (62, 80) | 71 (62, 81) | 0.700 |
| Resting SBP, median (IQR) | 133 (120, 148) | 136 (120, 150) | 132 (120, 148) | 0.020 |
| Resting DBP, median (IQR) | 80 (72, 84) | 80 (72, 84) | 80 (72, 85) | 0.13 |
| Stress Heart Rate, median (IQR) | 118 (92,144) | 113 (90, 137) | 119 (92, 144) | <0.001 |
| Stress SBP, median (IQR) | 150 (130, 170) | 150 (130, 170) | 150 (130, 170) | <0.001 |
| Stress DBP, median (IQR) | 80 (70, 84) | 80 (72, 84) | 80 (72, 85) | >0.900 |
| MPI Quantitative Imaging Analysis Parameters |  |  |  |  |
| Stress Ejection Fraction, median (IQR) | 63.9<br>(56.0, 71.2) | 56.9<br>(46.9, 64.4) | 64.2<br>(56.4, 71.5) | <0.001 |
| Ischemic TPD, median (IQR) | 2.1 (0.6, 4.6) | 7.2 (3.4, 11.8) | 2.0 (0.6, 4.4) | <0.001 |
| Rest TPD, median (IQR) | 0.1 (0.0, 2.0) | 1.7 (0.1, 6.6) | 0.1 (0.0, 1.9) | <0.001 |
| LV Volume, median (IQR) | 81.9<br>(63.2, 106.4) | 97.4<br>(75.6, 124.7) | 81.3<br>(62.7, 105.4) | <0.001 |
| LV Wall Volume, median (IQR) | 133.9<br>(115.8, 155.3) | 145.6<br>(126.7, 168.9) | 133.4<br>(115.4, 154.7) | <0.001 |
| Heart Shape Index, median (IQR) | 0.6 (0.5, 0.6) | 0.6 (0.5, 0.6) | 0.6 (0.5, 0.6) | <0.001 |
| Endpoint Events |  |  |  |  |
| Death or MI, n (%) | 4,323 (9.6%) | 279 (14.2%) | 4,044 (9.3%) | <0.001 |
| Death or MI follow-up period, year | 3.6 (2.7, 4.9) | 3.4 (2.2, 4.4) | 3.7 (2.7, 5.0) | <0.001 |

Values are presented as N (%) or median (IQ1, IQ3). Abbreviations: *BMI* body mass index, *CAD* coronary artery disease, *DBP* diastolic blood pressure, *IQR* interquartile range, *LV* left ventricle, *MI* myocardial infarction, *SBP* systolic blood pressure, *TPD* total perfusion deficit.

Supplementary Table 1 presents the characteristics of patients from the propensity-matched training dataset (n=2,010) and the held-out testing set (n=22,626). The flowchart of this study was presented in Supplementary Figure 3. Among the 22,626 participants in held-out testing, 9,927 (43.9%) were female, with a median age of 65 years (IQR 57–73). Of these, 958 (4.3%) underwent early revascularization, while the remaining patients received medical therapy.

### 3.2 Outcomes

During median follow-up of 3.51 years, 277 patients experienced death or MI in the propensity-matched training dataset. In the held-out testing set, 2,161 patients experienced death or MI during median follow-up of 3.7 years. The incidence of all-cause mortality or MI was significantly higher in the group that underwent early revascularization compared to the medical therapy group (14.2% vs. 9.3%, p<0.001), as shown in Table 1.

### 3.3 Predictive performance

The XGBoost model demonstrated strong predictive performance in forecasting 5-year all-cause mortality in the held-out test set (Supplementary Figure 4a). The AI model achieved an AUC of 0.76 (95% CI: 0.75–0.77), significantly outperforming ischemic TPD (p<0.001), which had an AUC of 0.59 (95% CI: 0.58–0.60). The top 10 features influencing the prediction included age, stress LVEF, stress diastolic blood pressure, peak stress heart rate, heart shape index, resting systolic blood pressure, resting heart rate, stress systolic blood pressure, LV wall volume, and peripheral vascular disease (Supplementary Figure 4b). Calibration plot of XGBoost model for 5-year all-cause mortality or MI is shown in Supplementary Figure 5.

### 3.4 Identifying Patients for Revascularization

In Figure 1, AI-recommended early revascularization (n=1,335; 5.9%) was associated with a significantly lower mortality or MI rate, demonstrating a 50% reduction in risk (adjusted HR 0.50, 95% CI: 0.25–1.00, p<0.05). By contrast, myocardial ischemia ≥10% (n=1,335; 5.9%) showed a numerically lower but non-significant trend (adjusted HR 0.59, 95% CI: 0.29–1.19, p=0.14, Supplementary Figure 6). Conversely, in patients for whom AI recommended medical therapy (n=21,291; 94.1%), there was an increased risk associated with early revascularization with a propensity-score-adjusted HR of 1.92 (95% CI: 1.52-2.44, p<0.001). In Supplementary Figure 6, patients with <10% myocardial ischemia (n=21,291; 94.1%), also had an increased risk of death or MI with early revascularization with a propensity-score-adjusted HR of 1.91 (95% CI: 1.50-2.44, p<0.001).

**Figure 1.**
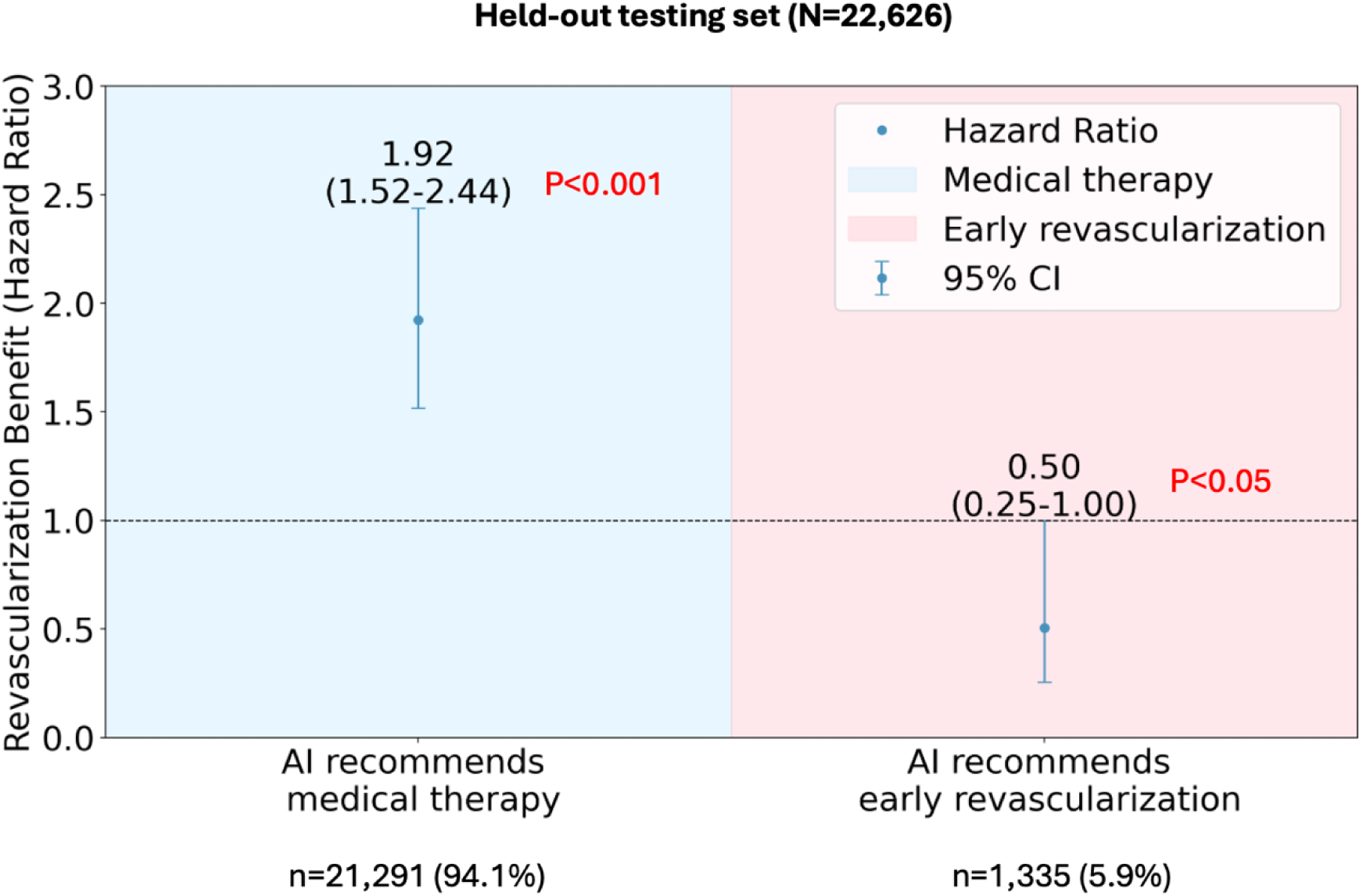
Risk of Early Revascularization: AI-selected patients. The AI model identified patients for early revascularization with a reduced associated risk (HR 0.50, 95% CI: 0.25–1.00, p<0.05). In contrast, patients recommended for medical therapy had an elevated risk of adverse outcomes (HR 1.92, 95% CI: 1.52–2.44, p<0.001). Hazard ratios were adjusted using propensity scores, which included age, sex, hypertension, dyslipidemia, peripheral vascular disease, smoking, presence of chest pain, prior myocardial infarction, family history of CAD, stress test type, ECG response to stress, stress LVEF, site, ischemic TPD, and rest TPD. Abbreviations: *AI* artificial intelligence, *CAD* coronary artery disease, *CI* confidence interval, *ECG* electrocardiograph, *LVEF* left ventricular ejection fraction, *TPD* total perfusion deficit.

In Figure 2, Kaplan-Meier curves show survival free of death or MI as a function of AI recommendation for revascularization and actual selection for early revascularization. Patients managed consistent with the AI recommendations had the best outcomes, while patients managed with a discordant strategy did worse (p<0.001). Patients in whom AI recommended an early invasive strategy but did not undergo early revascularization had the lowest survival rate (p<0.001), followed by patients managed with early revascularization when AI would recommend medical therapy.

**Figure 2.**
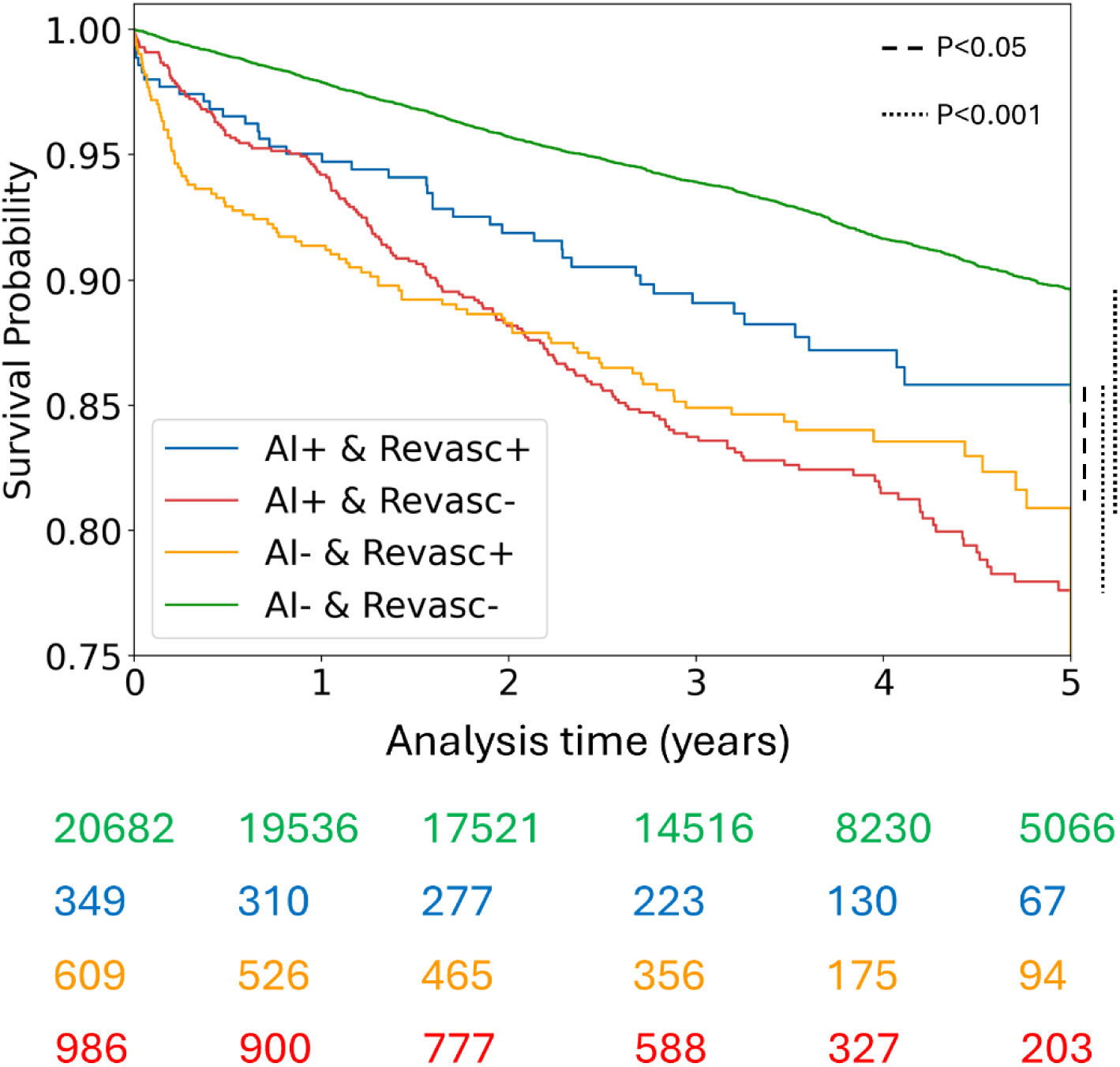
Kaplan-Meier Curve: AI Recommendations vs. Actual Revascularization. The numbers below the figure represent the number of patients at-risk at each time point. Patients whom the AI recommended for an invasive strategy but did not undergo early revascularization had the lowest survival rate by the end of the follow-up period. Notably, those who were not recommended for revascularization yet received the procedure also exhibited a declining survival rate over time. Abbreviations: *AI* artificial intelligence, *Revasc* early revascularization.

In Figure 3, a Venn diagram illustrates the overlap among patients recommended for revascularization by AI, those identified based on percent ischemia, and patients who actually underwent early revascularization. The AI model captured a larger proportion of patients who received revascularization compared to ischemia-based selection. Additionally, Table 2 presents a comparison of patient characteristics across these groups. Patients identified by AI had significantly lower LVEF, and a higher prevalence of hypertension, diabetes, and dyslipidemia (Figure 4).

**Figure 3:**
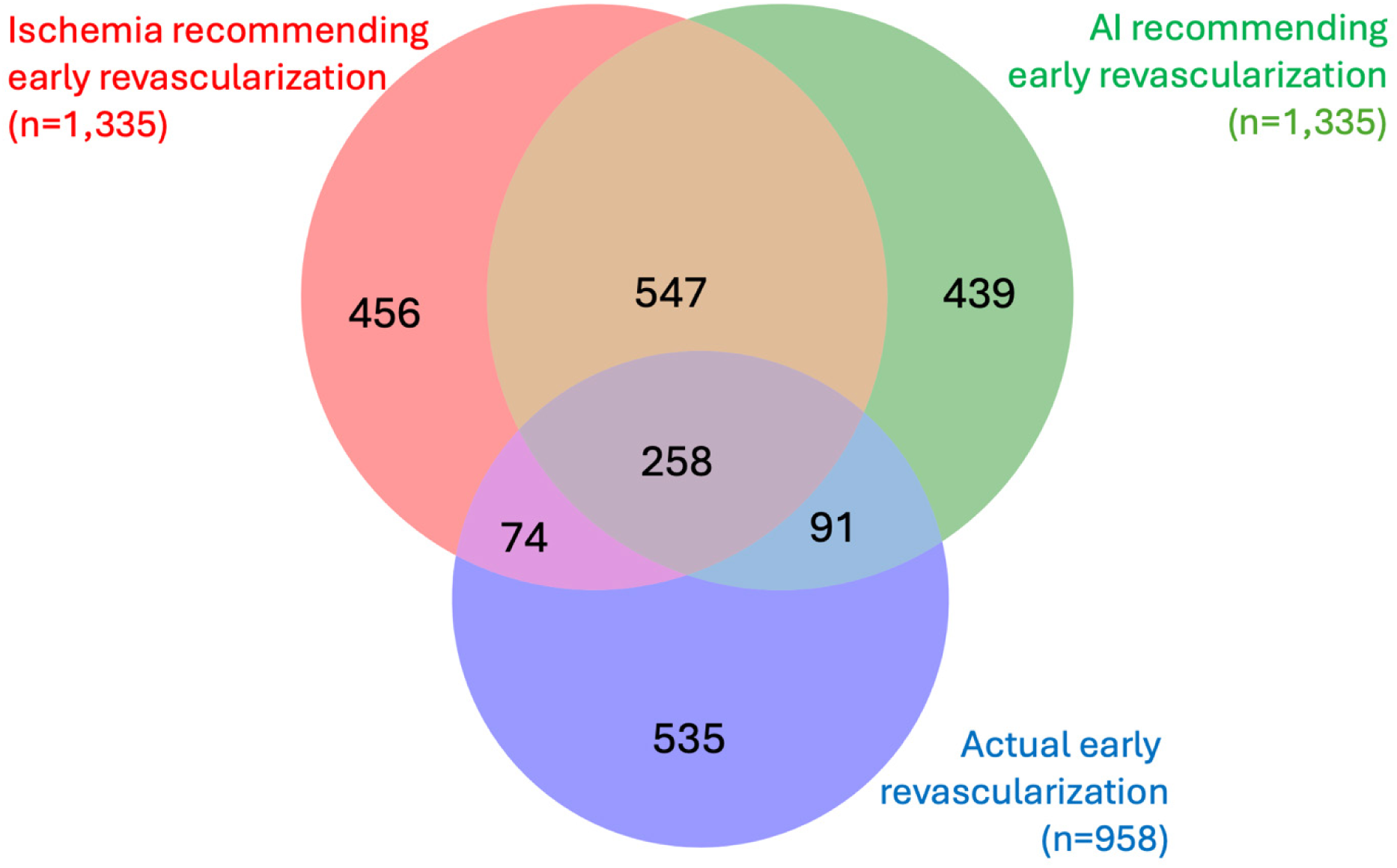
Venn Diagram: Ischemia, AI, and Actual Revascularization. This Venn diagram illustrates the overlap between ≥10% ischemia, AI-based predictions, and actual early revascularization. The red, green, and blue circles represent each category, with overlapping areas indicating agreement. The AI model captured a larger proportion of patients who received revascularization compared to ischemia-based selection. Abbreviations: *AI* artificial intelligence.

**Figure 4.**
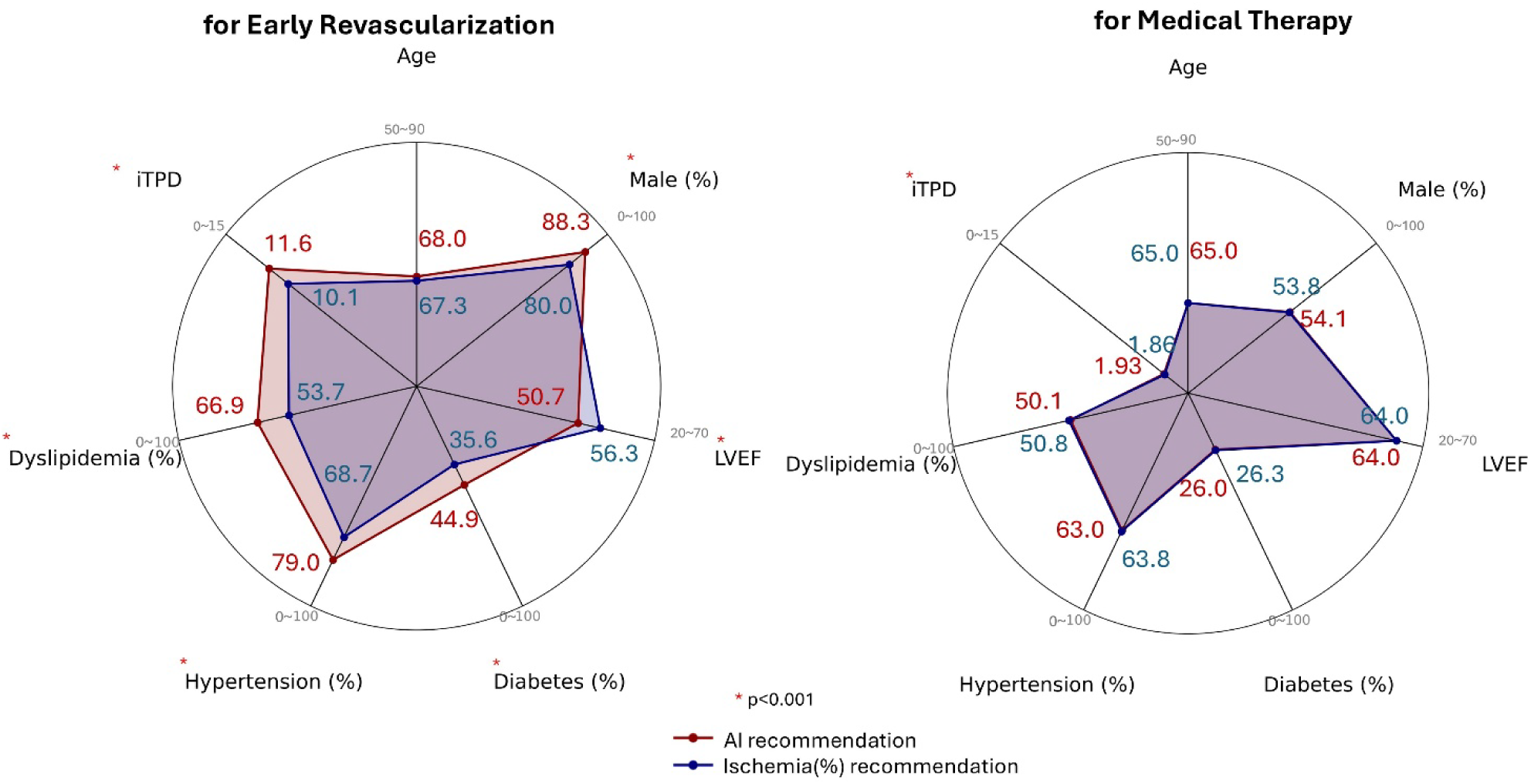
Radar Chart: Key Features Driving Treatment Recommendations. **Left panel:** Among patients recommended for an invasive strategy, those identified by AI had significantly lower LVEF, were older, and had a higher prevalence of hypertension, diabetes, and dyslipidemia. They also included a greater proportion of male patients and exhibited higher iTPD. **Right panel:** Among patients recommended for medical therapy, those identified by AI had a comparable prevalence of hypertension, diabetes, and dyslipidemia. However, they had a greater proportion of male patients and lower ischemia. Abbreviations: *AI* artificial intelligence, *iTPD* ischemic total perfusion deficit, *LVEF* left ventricle ejection fraction.

**Table 2.** Actual vs. AI vs. % Ischemia Revascularization Decisions in Held-Out Test Set (N=22,626)

|  | **Actual**<br>**Revascularization**<br>**n = 958** | **AI**<br>**n=1,335** | **% Ischemia**<br>**n=1,335** | **p-value** |
| --- | --- | --- | --- | --- |
| Age, median (IQR) | 66.0<br>(59.0, 73.2) | 68.0<br>(60.2, 75.0) | 68.0<br>(60.0, 75.4) | <0.001 |
| Male, n (%) | 746 (77.9%) | 1,179 (88.3%) | 1,059 (79.4%) | <0.001 |
| **Past Medical History** |  |  |  |  |
| Hypertension, n (%) | 699 (73.0%) | 1,054 (79.0%) | 945 (71.2%) | <0.001 |
| Diabetes Mellitus, n (%) | 366 (38.2%) | 599 (44.9%) | 495 (37.1%) | <0.001 |
| Hyperlipidemia, n (%) | 547 (57.9%) | 892 (66.9%) | 788 (59.5%) | <0.001 |
| Family History of CAD, n (%) | 328 (34.2%) | 472 (35.4%) | 357 (26.9%) | <0.001 |
| Peripheral Vascular Disease, n (%) | 313 (33.1%) | 596 (45.9%) | 335 (27.3%) | <0.001 |
| Smoking, n (%) | 264 (27.6%) | 394 (29.5%) | 337 (25.3%) | 0.048 |
| **MPI Acquisition Parameters** |  |  |  |  |
| Exercise Stress, n (%) | 414 (44.0%) | 525 (39.6%) | 533 (40.8%) | 0.106 |
| Resting Heart Rate, median (IQR) | 71 (62, 81) | 68 (60, 78) | 70.00<br>(62.00, 80.00) | <0.001 |
| Resting BP Systole, median (IQR) | 134 (122, 150) | 135 (120, 149) | 135.00<br>(121.00, 150.00) | 0.801 |
| Resting BP Diastole, median (IQR) | 80 (72, 84) | 80 (70, 84) | 80.00<br>(72.00, 84.00) | 0.024 |
| Stress Heart Rate Peak (Pharm/Ex), median (IQR) | 114 (91, 137) | 105 (86, 133) | 105.00<br>(88.00, 133.00) | <0.001 |
| Stress Systolic BP Peak (Pharm/Ex), median (IQR) | 150 (130, 170) | 142 (124, 162) | 140.50<br>(125.00, 160.00) | 0.001 |
| Stress Diastolic BP Peak (Pharm/Ex), median (IQR) | 80 (70, 83) | 80 (70, 80) | 80.00<br>(70.00, 82.00) | 0.008 |
| **MPI Quantitative Imaging Analysis Parameters** |  |  |  |  |
| Stress Ejection Fraction, median (IQR) | 56.4<br>(46.3, 64.1) | 50.7<br>(38.5, 59.3) | 53.63<br>(42.86, 63.14) | <0.001 |
| Ischemic TPD, median (IQR) | 7.5<br>(3.6, 12.0) | 11.6<br>(7.8, 16.1) | 13.04<br>(11.25, 16.30) | <0.001 |
| Rest TPD, median (IQR) | 1.8 (0.1, 7.1) | 4.0 (0.5, 11.2) | 3.44<br>(0.49, 9.08) | <0.001 |
| LV Volume, median (IQR) | 99.3<br>(75.6, 127.8) | 112.5<br>(84.8, 149.2) | 104.75<br>(76.57, 137.64) | <0.001 |
| LV Wall Volume, median (IQR) | 147.4<br>(127.2, 170.7) | 157.0<br>(135.4, 186.2) | 150.74<br>(129.23, 179.29) | <0.001 |
| Heart Shape Index, median (IQR) | 0.6 (0.6, 0.6) | 0.6 (0.6, 0.7) | 0.61 (0.56, 0.67) | <0.001 |
| Endpoint Events |  |  |  |  |
| Death or MI, n (%) | 141 (14.7%) | 229 (17.2%) | 229 (17.2%) | 0.218 |

Values are presented as N (%) or median (IQ1, IQ3). Abbreviations: *AI* artificial intelligence, *BMI* body mass index, *BP* blood pressure, *CAD* coronary artery disease, *Ex* exercise testing, *IQR* interquartile range, *LV* left ventricle, *MI* myocardial infarction, *Pharm* pharmacological testing, *TPD* total perfusion deficit.

## 4. DISCUSSION

We evaluated whether AI could be leveraged to improve the selection of patients for revascularization using data from a large, multicenter, international registry population. We used a novel digital twin approach to simulate each patient’s outcome when managed with or without early revascularization. By simulating both management strategies the framework computed individualized treatment effects that would otherwise be unobservable^23^. Importantly, we demonstrated that patients who would be selected for early revascularization by AI-REVASC had a significant reduction in death or MI when managed with early revascularization, while those selected by ischemia alone did not. This personalized approach for decision making – based on expected benefit from a therapy and instead of overall risk – may finally allow physicians to accurately identify patients with chronic coronary syndromes who derive a survival benefit from revascularization.

While treatment benefit is traditionally evaluated by randomized controlled trials (RCT), they have well-documented limitations in evaluating individualized treatment benefit. RCTs often exclude high-risk or heterogeneous subpopulations (e.g., patients with severely reduced LVEF or complex comorbidities), thereby limiting external validity. Moreover, RCTs are constrained by logistical, and economic considerations that preclude the exhaustive testing of all potential treatment strategies in all patient types. In contrast, by training on propensity-matched data, the model mimics RCT-like covariate balance while preserving real-world heterogeneity. While not a replacement for RCTs, this framework can simulate RCTs *in silico* to inform trial design and guide therapy in diverse clinical populations.

A critical question about any imaging test is whether it can be used to guide patient management to improve outcomes. In this regard, most patients with evidence of coronary atherosclerosis (based on coronary artery calcification^24, 25^ or perfusion abnormalities^26, 27^) may benefit from initiation of medical therapy. However, a more difficult clinical challenge is identifying which patients with chronic coronary syndromes may benefit from early revascularization. Large observational studies suggested that patients with moderate to severe ischemia may benefit from early revascularization^4–9^. However, the ISCHEMIA trial failed to show improvement in the primary outcome (cardiovascular death, MI, hospitalization for unstable angina, heart failure, or resuscitated cardiac arrest) during a median follow-up of 3.2 years^28^ and no difference in all-cause mortality at 7 years^29^. In our work, patients who received management concordant with the AI recommendation had the lowest risk of death or MI. Meanwhile, patients managed with early revascularization when AI recommended medical therapy had the highest risk. Furthermore, the predicted benefit from early revascularization was greater for patients selected using AI compared to patients selected based on ischemia alone. Overall, these findings suggest that AI could support physician decision-making when deciding which patients should be referred for revascularization after MPI.

The discrepancy between observational and randomized studies suggests that ischemia alone is inadequate in selecting patients for revascularization. One possibility for this discordance is that patients excluded from the ISCHEMIA trial are those most likely to benefit^30^. For example, patients with very reduced LVEF were excluded from the ISCHEMIA trial but may derive benefit from revascularization in the presence of less extensive ischemia compared to patients with preserved ventricular function^7^. This is consistent with the recommendations for revascularization by AI in the present study, where LVEF was one of the most important features. Observational studies have demonstrated that many different features may influence the relationship between ischemia and benefit from revascularization. Patients with diabetes (which was also a critical feature in the present study) may benefit from revascularization in the presence of less extensive ischemia^12^, while patients with prior CAD may only derive benefit when more significant ischemia is present^9^. The total number of features that could be considered makes it difficult to apply clinically in a systematic fashion. Lastly, it is critical to consider that overall risk may not be directly related to therapeutic benefit from revascularization. A decision support tool, such as the proposed AI-REVASC score, based on a comprehensive assessment of expected benefit (rather than risk alone) could potentially dramatically improve selection of patients for revascularization.

Clinical selection of patients for revascularization has always relied on factors influencing perceived risk and benefit beyond ischemia. As a reflection of this, less than 1 in 5 patients with ischemia ≥10% in our population, actually underwent revascularization. This selection bias is an important consideration which was incorporated in our analysis through the use of propensity-matched analyses (in the training population) and propensity score adjustment in the held-out testing population. Compared to ischemia-based selection, there was closer agreement between AI-based selection and actual clinical practice. Patients who were managed with concordant treatment strategies had the lowest risk of death or MI. Meanwhile, patients who underwent revascularization when AI recommended medical therapy were at the highest risk.

This may reflect improved selection with AI or residual unmeasured confounding features (such as refractory symptoms or known high risk coronary anatomy) which could drive revascularization decisions and are associated with adverse outcomes^31^. Prospective randomized studies would be needed to discern these two possibilities and are warranted given the potential clinical utility of AI-based decision support tools.

We recognized that our computational model, while simpler than full-fledged industrial digital twin systems, remained sufficient to estimate potential risks under alternative treatment scenarios. Importantly, the medical community has yet to establish a consensus definition of what constitutes a digital twin in healthcare ^23^. Our work contributes meaningful empirical evidence to this evolving field. Collectively, our AI approach not only strengthens the validity of the effect estimates but also empowers the AI algorithm to uncover heterogeneous responses across patient subgroups**—**advancing precision medicine in cardiac care.

### Study Limitations

Our study has a few important limitations. The study was observational and retrospective. All-cause mortality or MI rather than cardiac death was used in our outcome. While cardiac death may be more closely related than all-cause mortality to MPI findings and revascularization, collecting data relating to cardiac death was not possible within the context of this large, multicenter registry, as in most of the sites, the databases did not differentiate causes of death. We also cannot exclude that our results are related to residual confounders. Prospective studies are needed to validate the clinical utility of this AI approach for revascularization selection.

## Conclusion

AI integrates diverse clinical, imaging, and demographic factors beyond ischemia to enhance patient selection for early revascularization after MPI. This study examines whether AI can improve identification of patients who benefit from revascularization. Our findings demonstrate that the AI model effectively stratifies patients, highlighting those more likely to experience improved outcomes with early intervention. By leveraging AI-driven insights, this approach has the potential to refine clinical decision-making and optimize treatment strategies.

## Data Availability

To the extent allowed by data and code sharing agreements and IRB protocols, the deidentified data and analysis code from this manuscript will be shared upon written request. The analysis code will be made available on GitHub: https://github.com/qimagingAI/AI4RevascBenefit/.

## Acknowledgements

This research was supported in part by grants R01HL089765 and R35HL161195 from the National Heart, Lung, and Blood Institute at the National Institutes of Health (PI: Piotr Slomka) and a grant from the Adelson Medical Research Foundation. The content is solely the responsibility of the authors and does not necessarily represent the official views of the National Institutes of Health.

## Contributors

WZ conducted algorithm development, data processing, experiments, analysis, and co-wrote the manuscript. RJM co-wrote the manuscript and contributed to materials, algorithm development, and clinical expertise. PJS designed the study, provided overall guidance and study funding, co-wrote the manuscript, and contributed materials, clinical expertise, and technical expertise. AJE, AJS, AS, DD, DSB, EM, GR, JXL, JY, JZ, JH, KP, MF, MFDC, ML, MM, PBK, PK, PC, RJM, SD, TB, TH, TR, TS, VB, and WA contributed to materials, clinical expertise, and technical expertise. All authors critically revised the manuscript and contributed to its formation. WZ, GR, RJM, ML, and PJS directly accessed and verified the data in the study. All authors had full access to all data in the study, accept the final responsibility for submission, and take responsibility for the contents of the manuscript.

## ABBREVIATIONS

AI: artificial intelligence
CABG: coronary artery bypass grafting
CAD: coronary artery disease
ISCHEMIA: International Study of Comparative Health Effectiveness with Medical and Invasive Approaches
LVEF: left ventricular ejection fraction
MI: myocardial infarction
MPI: myocardial perfusion imaging
PCI: percutaneous coronary intervention
SPECT: single photon emission computed tomography
TPD: total perfusion deficit

