## Supplementary Materials for "AI-based identification of patients who benefit from revascularization: a multicenter study"

### Supplementary Tables

**Supplementary Table 1: Baseline patient characteristics by development set.**

| | **Propensity matched Training n = 2,010 (4.3 %)** | **Testing n=22,626 (95.7%)** | **p-value** |
|---|---|---|---|
| Age, median (IQR) | 67.0 (59.0, 74.7) | 65.0 (57.0, 73.0) | <0.001 |
| Male, n (%) | 1,537 (76.5%) | 12,699 (56.1%) | <0.001 |
| BMI, median (IQR) | 28.4 (25.3, 32.2) | 28.4 (25.1, 32.6) | 0.7 |
| Initial treatment, n (%) | | | <0.001 |
| Early revascularization | 1,005 (50%) | 958 (4.3%) | |
| Medical Therapy | 1,005 (50%) | 21,668 (95.7%) | |
| Race, n (%) | | | <0.001 |
| American Indian or Alaska Native | 14 (0.7%) | 64 (0.3%) | |
| Asian | 25 (1.2%) | 243 (1.1%) | |
| Black or African American | 74 (3.7%) | 1,267 (5.6%) | |
| Native Hawaiian or Other Pacific Islander | 0 (0.0%) | 18 (0.1%) | |
| White | 651 (32.4%) | 6,948 (30.7%) | |
| Not available | 1,246 (62.0%) | 14,086 (62.3%) | |
| Past Medical History | | | |
| Past CAD, n (%) | 439 (21.8%) | 2,650 (11.7%) | <0.001 |
| Hypertension, n (%) | 1,470 (73.1%) | 14,486 (64.0%) | <0.001 |

| | | | |
|---|---|---|---|
| Diabetes Mellitus, n (%) | 784 (39.0%) | 6,126 (27.1%) | < 0.001 |
| Hyperlipidemia, n (%) | 1,207 (60.0%) | 11,474 (50.7%) | <0.001 |
| Family History of CAD, n (%) | 690 (34.3%) | 7,062 (31.2%) | 0.004 |
| Peripheral Vascular Disease, n (%) | 717 (35.7%) | 4,287 (18.9%) | <0.001 |
| Smoking, n (%) | 584 (29.1%) | 5,431 (24.0%) | <0.001 |
| MPI Acquisition Parameters | | | |
| Exercise Stress, n (%) | 762 (37.9%) | 10,551 (46.6%) | <0.001 |
| Resting Heart Rate, median (IQR) | 70 (62, 80) | 71 (62, 81) | 0.041 |
| Resting BP Systole, median (IQR) | 134 (120, 150) | 132 (120, 148) | 0.074 |
| Resting BP Diastole, median (IQR) | 80 (72, 84) | 80 (72, 84) | 0.005 |
| Stress Heart Rate Peak (Pharm/Ex) , median (IQR) | 110 (88, 136) | 119 (92, 144) | <0.001 |
| Stress Systolic BP Peak (Pharm/Ex) , median (IQR) | 145 (124, 170) | 150 (130, 170) | <0.001 |
| Stress Diastolic BP Peak (Pharm/Ex) , median (IQR) | 80 (70, 83) | 80 (70, 84) | 0.009 |
| MPI Quantitative Imaging Analysis Parameters | | | |
| Stress Ejection Fraction, median (IQR) | 56.70 (46.16, 64.73) | 63.81 (55.88, 71.22) | <0.001 |
| Ischemic TPD, median (IQR) | 6.37 (2.95, 11.26) | 2.13 (0.63, 4.65) | <0.001 |
| Rest TPD, median (IQR) | 1.57 (0, 6.57) | 0.09 (0, 2.05) | <0.001 |
| LV Volume, median (IQR) | 98.05 (75.44, 126.14) | 82.28 (63.24, 106.92) | <0.001 |
| LV Wall Volume, median (IQR) | 144.83 (126.38, 170.72) | 134.20 (115.93, 155.66) | <0.001 |
| Heart Shape Index, median (IQR) | 0.59 (0.55, 0.65) | 0.58 (0.53, 0.62) | <0.001 |
| Endpoint Events | | | |
| Death or MI, n(%) | 277 (13.8%) | 2,161 (9.6%) | <0.001 |

| | | | |
|---|---|---|---|
| Death or MI follow-up period, year | 3.51 (2.49, 4.54) | 3.65 (2.69, 4.93) | <0.001 |

Values are presented as N (%) or median (IQ1, IQ3).

*BMI* body mass index, *BP* blood pressure, *CABG* coronary artery bypass graft, *CAD* coronary artery disease, *Ex* exercise testing, *IQR* interquartile range, *LV* left ventricle, *MI* myocardial infraction, *Pharm* pharmacological testing, *SDS* summed difference score, *TPD* total perfusion deficit.

**Supplementary Figures**

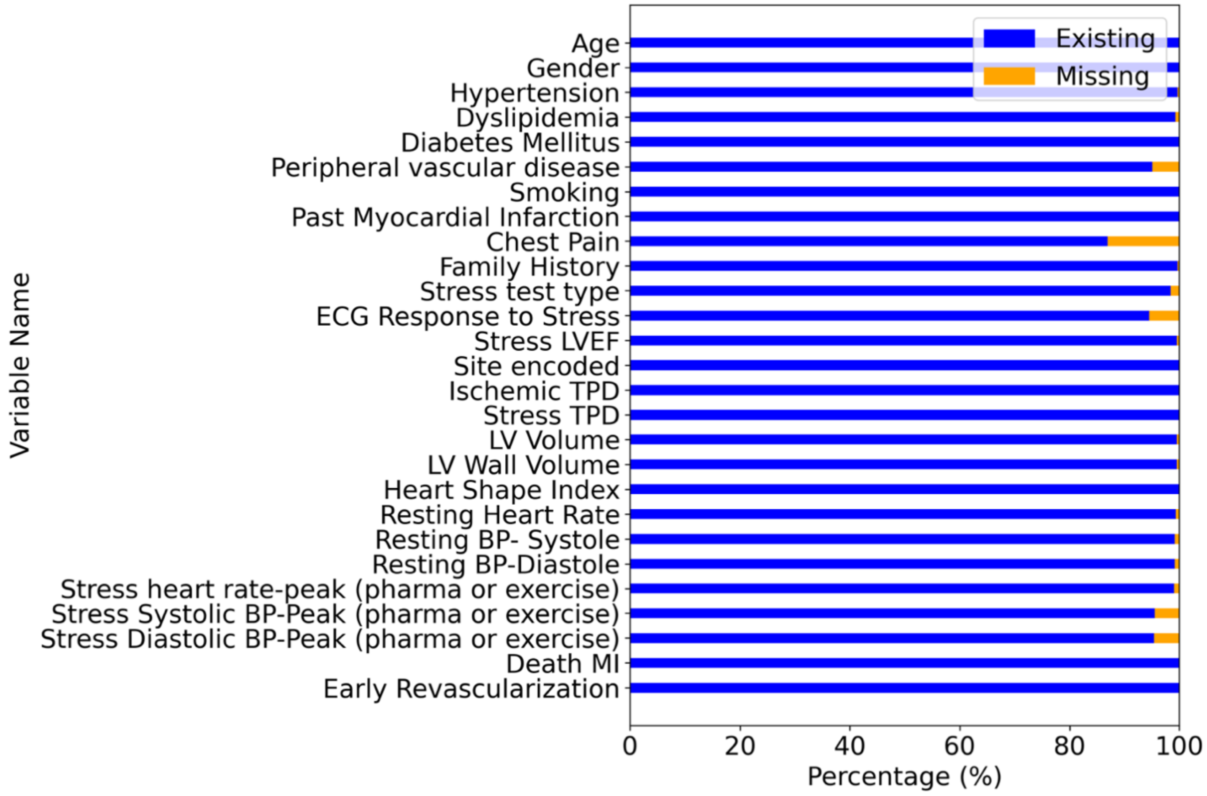


**Supplementary Figure 1: The percentage (%) of missingness of variables included in the AI model.** All the variables have missingness less than 20%. No data imputation was utilized as XGBoost handles missing values internally. Abbreviations: *BP* blood pressure, *ECG* electrocardiogram*, LV* left ventricle, *LVEF* left ventricle ejection fraction, *MI* myocardial infraction, *TPD* total perfusion deficit, *XGBoost* extreme gradient boosting.

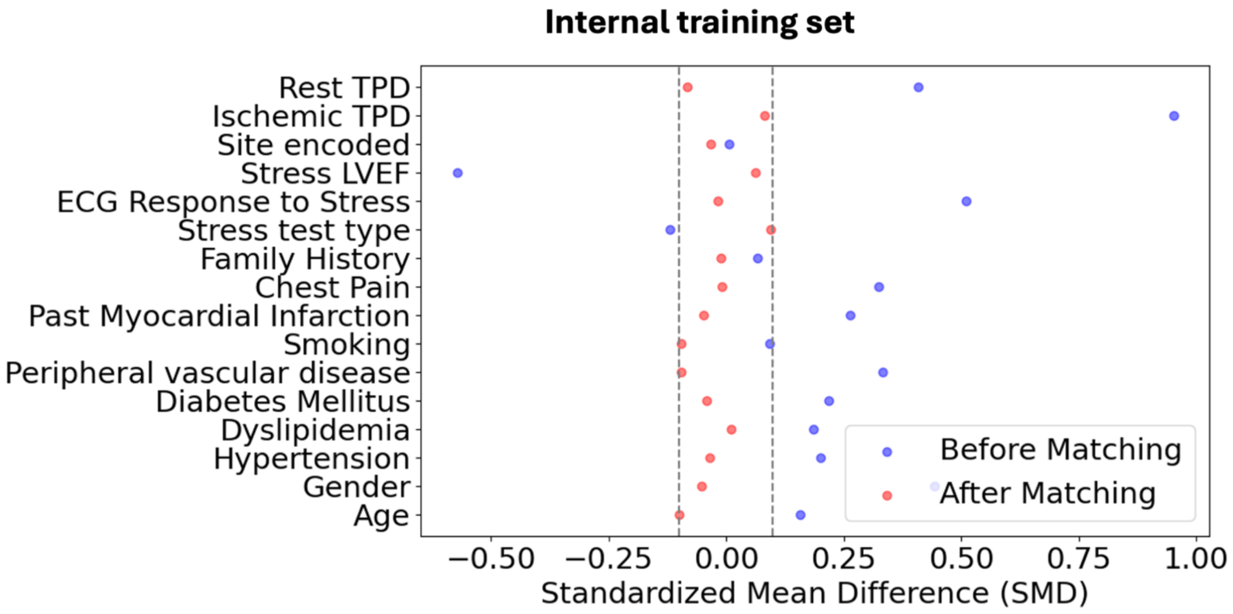


**Supplementary Figure 2: Absolute standardized differences after adjustment for propensity score for training and testing set.** Adjusted differences >0.100 (dashed black line) are considered significantly different. Abbreviations: *ECG* electrocardiogram*, LVEF* left ventricle ejection fraction, *TPD* total perfusion deficit.

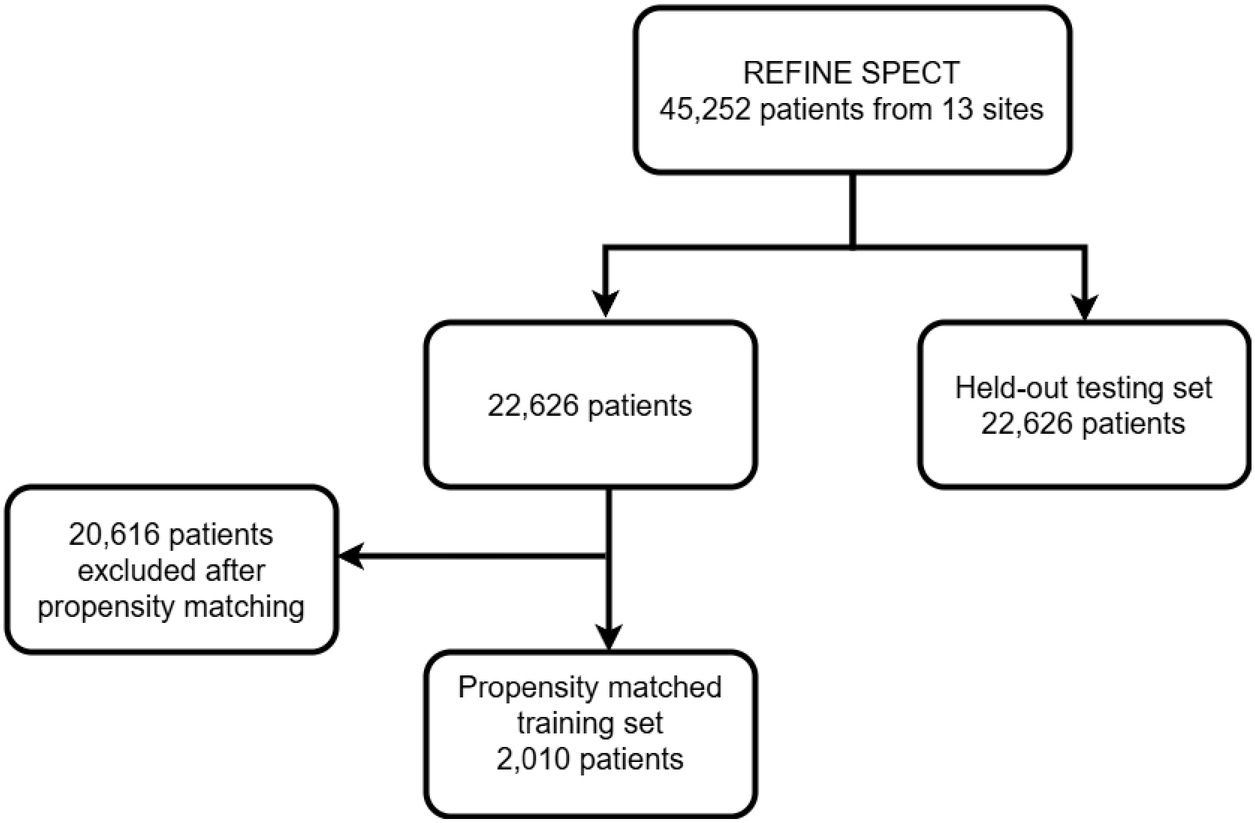


**Supplementary Figure 3: Flowchart of the study.** The dataset (N=45,252) was randomly and evenly divided into two parts. The first part was applied with propensity matching to simulate a randomized control trial with 1:1 matching on early revascularization and medical therapy. 20,616 patients who received medical therapy were not matched and were excluded from training. The AI model was developed on the propensity matched training set which contains 2,010 patients. Model was evaluated on the held-out testing set with 22,626 patients.

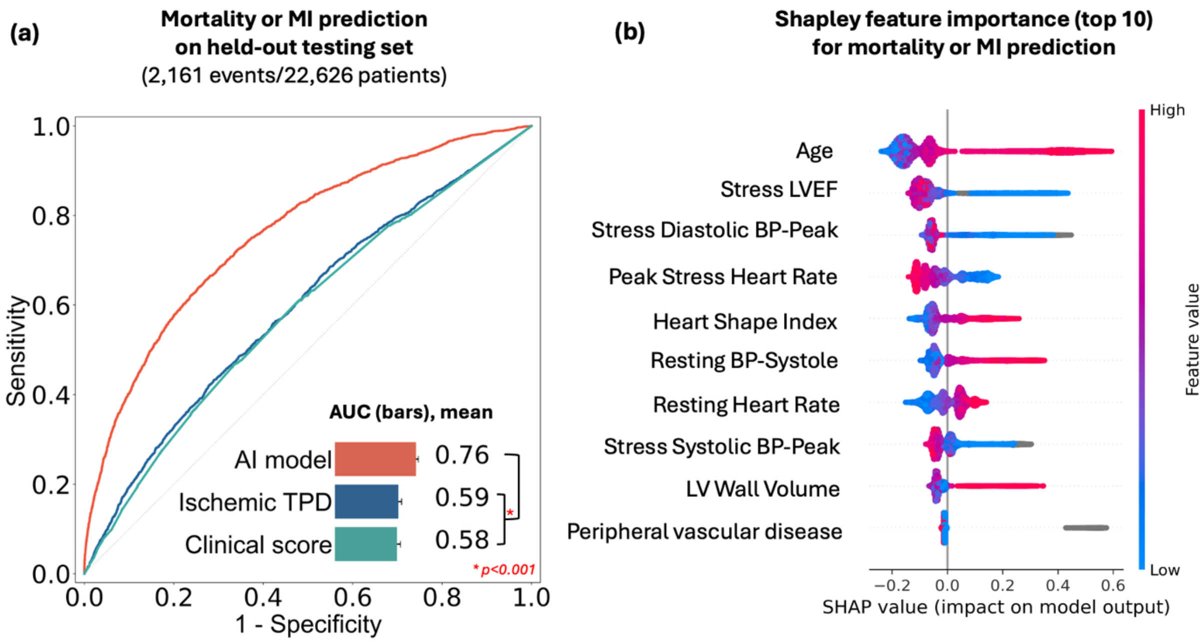


**Supplementary Figure 4: Prediction performance from held-out testing. (a)**: Receiver-operating characteristics curve for all-cause mortality or myocardial infarction (MI) and areas under the curve (AUC) values. AI model incorporates age, sex, past medical therapy, stress test type, rest and stress vitals, ischemia total perfusion deficit (TPD), ejection fractions, shape index, left ventricle volume, transient ischemic dilation. **(b):** Shapley feature importance (top 10) for mortality or MI prediction. Each dot represents an individual patient and a specific feature. The distance of each dot from the center line (0 on the x-axis) indicates how strongly that feature drives the prediction, either positively or negatively, for that patient.

Abbreviations: *BP* blood pressure, *LV* left ventricle, *LVEF* left ventricle ejection fraction, *SHAP* shapley additive explanations.

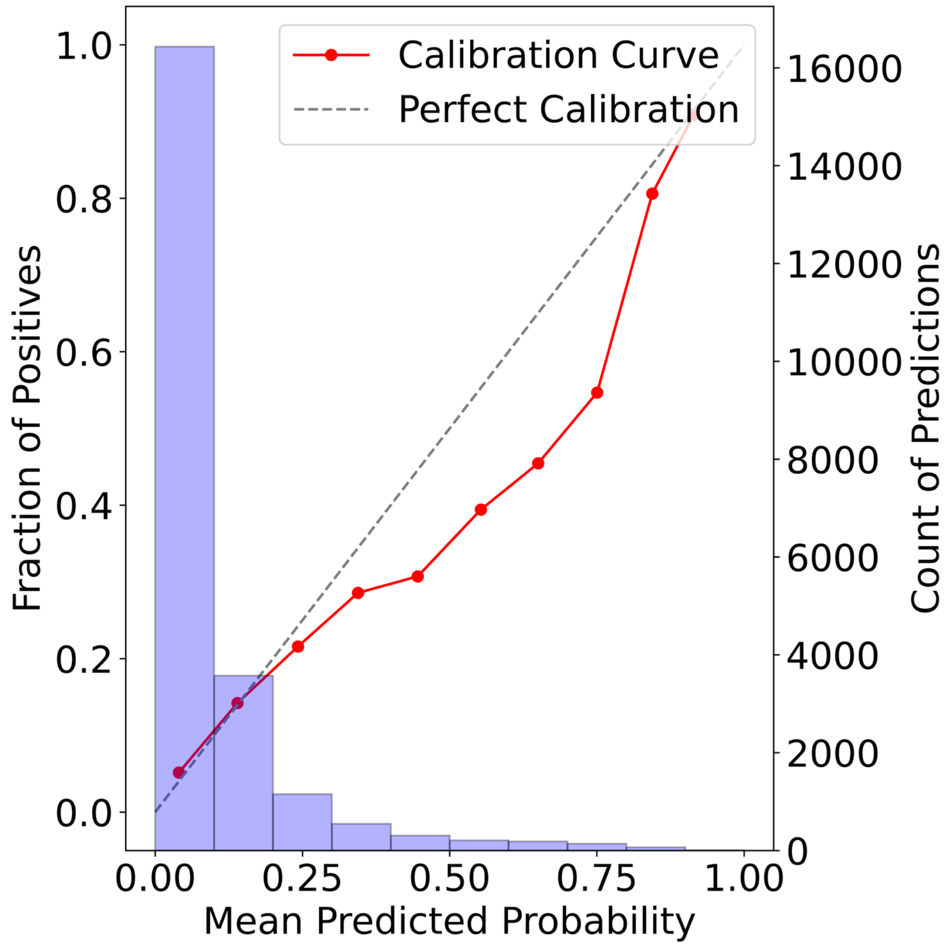


**Supplementary Figure 5: Calibration plots for 5-year all-cause mortality and myocardial infarction (MI) in held-out testing.** Abbreviations: *XGBoost* extreme gradient boosting.

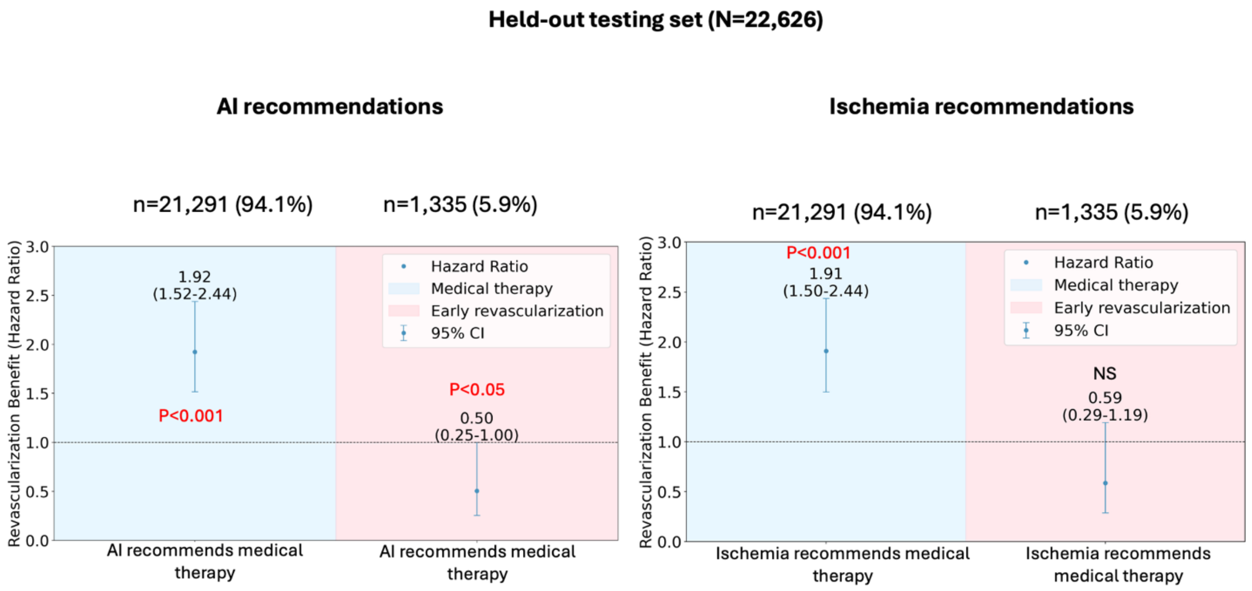


**Supplementary Figure 6. Risk of Early Revascularization: AI vs. Ischemia-Based Selection.** Patients selected by the AI model showed lower associated risk compared to those selected by ischemia in the held-out testing set. The hazard ratios for medical therapy recommendations from both methods were comparable. Hazard ratios were adjusted by age, sex, hypertension, dyslipidemia, peripheral vascular disease, smoking, presence of chest pain, prior myocardial infarction, family history of CAD, stress test type, ECG response to stress, stress LVEF, site, ischemic TPD, and rest TPD.

Abbreviations: *AI* artificial intelligence, *CAD* coronary artery disease, *CI* confidence interval, *ECG* electrocardiograph, *LVEF* left ventricular ejection fraction, *NS* not significant, *TPD* total perfusion deficit.
